# Multiple introductions and country-wide spread of DENV-2 genotype II (Cosmopolitan) in Brazil

**DOI:** 10.1101/2023.06.06.23290889

**Authors:** Tiago Gräf, Caroline do Nascimento Ferreira, Gustavo Barbosa de Lima, Raul Emídio de Lima, Lais Ceschini Machado, Tulio de Lima Campos, Michelle Orane Schemberger, Helisson Faoro, Marcelo Henrique Santos Paiva, Matheus Filgueira Bezerra, Valdinete Nascimento, Victor Souza, Fernanda Nascimento, Matilde Mejía, Dejanane Silva, Yasmin Silva de Oliveira, Luciana Gonçalves, Tatyana Costa Amorim Ramos, Daniel Barros de Castro, Ana Ruth Arcanjo, LACEN-AM team, Herton Augusto Pinheiro Dantas, LAFRON-AM team, Mayra Marinho Presibella, LACEN-PR team, Sandra Bianchini Fernandes, LACEN-SC team, Tatiana Schaffer Gregianini, CEVS-RS team, Keilla Maria Paz e Silva, LACEN-PE team, Claudio Tavares Sacchi, IAL team, Ana Cecília Ribeiro Cruz, IEC team, Claudia Nunes Duarte dos Santos, Ana Maria Bispo de Filippis, Richard Steiner Salvato, Gonzalo Bello, Gabriel Luz Wallau, Felipe Naveca

**Author notes:** Lists of authors and their affiliations appear at the Supplementary Consortia List (LACEN-AM team, LAFRON-AM team, LACEN-PR team, LACEN-SC team, CEVS-RS team, LACEN-PE team, IAL team, IEC team).

## Abstract

Dengue virus serotype 2, genotype Cosmopolitan (DENV-2-GII), is one of the most widespread DENV strains globally. In the Americas, DENV-2 epidemics has been dominated by DENV-2 genotype Asian-American (DENV-2-GIII) and the first cases of DENV-2-GII were only described in 2019, in Peru, and in 2021 in Brazil. To gain new information about the circulation of DENV-2-GII in Brazil, we sequenced 237 DENV-2 confirmed cases sampled between March 2021 and March 2023 and revealed that DENV-2-GII is already present in all geographic regions of Brazil. Phylogeographic analysis inferred that DENV-2-GII was introduced at least four times in Brazil, between May 2020 and August 2022, generating multiple clades that spread throughout the country with different success. Despite multiple introductions of DENV-2-GII, analysis of the country-wide laboratory surveillance data showed that the Brazilian dengue epidemic in 2022 was dominated by DENV-1 in most states. We suggest that massive circulation of DENV-2-GIII in previous years in Brazil might have created a population immune barrier for widespread transmission of DENV-2-GII, leading to sustained cryptic circulation and localized outbreaks of this new genotype. In summary, our study stresses the importance of arboviral genomic surveillance to close monitoring and better understand the potential impact of DENV-2-GII in the coming years.

## Research letter

Dengue virus serotype 2 (DENV-2) can be divided into five endemic/epidemic (nonsylvatic) genotypes: American, Cosmopolitan, Asian-American, Asian-II, and Asian-I ^1^, also known as genotypes I-V, respectively. In the Americas, the DENV-2 American genotype (DENV-2-GI) was the first to be identified, in the 1940s. However, it was later replaced by the Asian-American genotype (DENV-2-GIII) in the 1980s, and cases of DENV-2-GI were not found since then^2^. In Brazil, DENV-2-GIII was first introduced in the country in the 1990s and at least three other introductions happened between 1990 and 2014, all with origin in Caribbean or Northern South America countries^3,4^. Besides DENV-2, in Brazil all four DENV serotypes were already identified, presenting a complex dynamics of introductions, co-circulation, and alternation of dominance, which is associated with an increasing number of severe dengue cases and deaths in recent years ^5^.

In 2021, the first cases of DENV-2 Cosmopolitan genotype (DENV-2-GII) were identified in the Brazilian states of Goiás (GO)^6^ and Acre (AC)^7^, located in the Central-Western and Northern country regions, respectively. Phylogenetic analysis suggested that the Brazilian DENV-2-GII sequences derived from an outbreak of this genotype in Peru in 2019^8^, representing the first documented introduction of DENV-2-GII in the Americas. DENV-2-GII is one of the most widespread genotypes, circulating in Asia-Pacific, Middle East, Africa, and Oceania, substantially contributing to the global dengue burden^9^. Here we present new data on the DENV-2-GII spread in Brazil, showing that shortly after multiple introductions in the country, it is already present in all geographical regions.

Genomic surveillance for DENV was performed in the Brazilian states of Amazonas (AM), Pernambuco (PE), São Paulo (SP), Paraná (PR), Santa Catarina (SC), and Rio Grande do Sul (RS) by each State Central Laboratories (LACEN) and Fiocruz laboratories. A selection of DENV-2 positive samples in RT-qPCR test were submitted for whole genome amplification and sequencing using Illumina’s Viral Surveillance Panel or COVIDseq Test adapted to DENV-2. Genomes were assembled in ViralFlow software^10^ or using Geneious Prime 2022 and consensus sequences were genotyped using online tools (methods detailed in **Supplementary text**).

Among 237 DENV-2 sequenced samples, we identified 60 DENV-2-GII (**Supplementary Table 1**) which were aligned with a DENV-2-GII representative global dataset. Maximum likelihood phylogenetic analysis revealed that all DENV-2-GII Brazilian genomes, from this and previous studies ^6,7^, clustered in a monophyletic clade with sequences from Peru and Bangladesh (**Supplementary Figure 1**). Our phylogeographic analysis estimated that DENV-2-GII was introduced from Bangladesh into Peru in April 2019 (November 2018 - August 2019, 95% High Posterior Density [HPD]), and from there, it was introduced in Brazil at least four times (**Figure 1A**). The first introduction occurred through the AC state (North) in May 2020 (June 2019 - November 2020, 95% HPD) and from AC, DENV-2-GII disseminated to all other four Brazilian regions represented by the states of GO (Central-West), PE (Northeast), SP (Southeast), and SC (South). The oldest DENV-2-GII Brazilian samples collected in 2021 in AC (February to March), PE (July) and GO (November) clustered within this clade. A second introduction of DENV-2-GII happened in the state of SP in March 2021 (September 2020 - July 2021, 95% HPD), and from there, DENV-2-GII spread southwards to the states of PR, SC, and RS. A third introduction of DENV-2-GII occurred in January 2022 (October 2021 - March 2022, 95% HPD) in the state of RS, without evidence of further dissemination to other Brazilian states. Finally, the most recent introduction happened in AM state in August 2022 (June 2022 - October 2022, 95% HPD), causing an outbreak in cities close to the border with Peru and Colombia^11^. It is interesting to note that the first introductions of DENV-2-GII into Brazil most probably occurred during the period (May 2020 - March 2021) in which the country was most heavily affected by the SARS-CoV-2 pandemic, and that despite the control measures implemented to restrict human mobility in that period, DENV-2-GII viruses introduced in AC and SP were able to disseminate to other Brazilian states.

**Figure 1.**
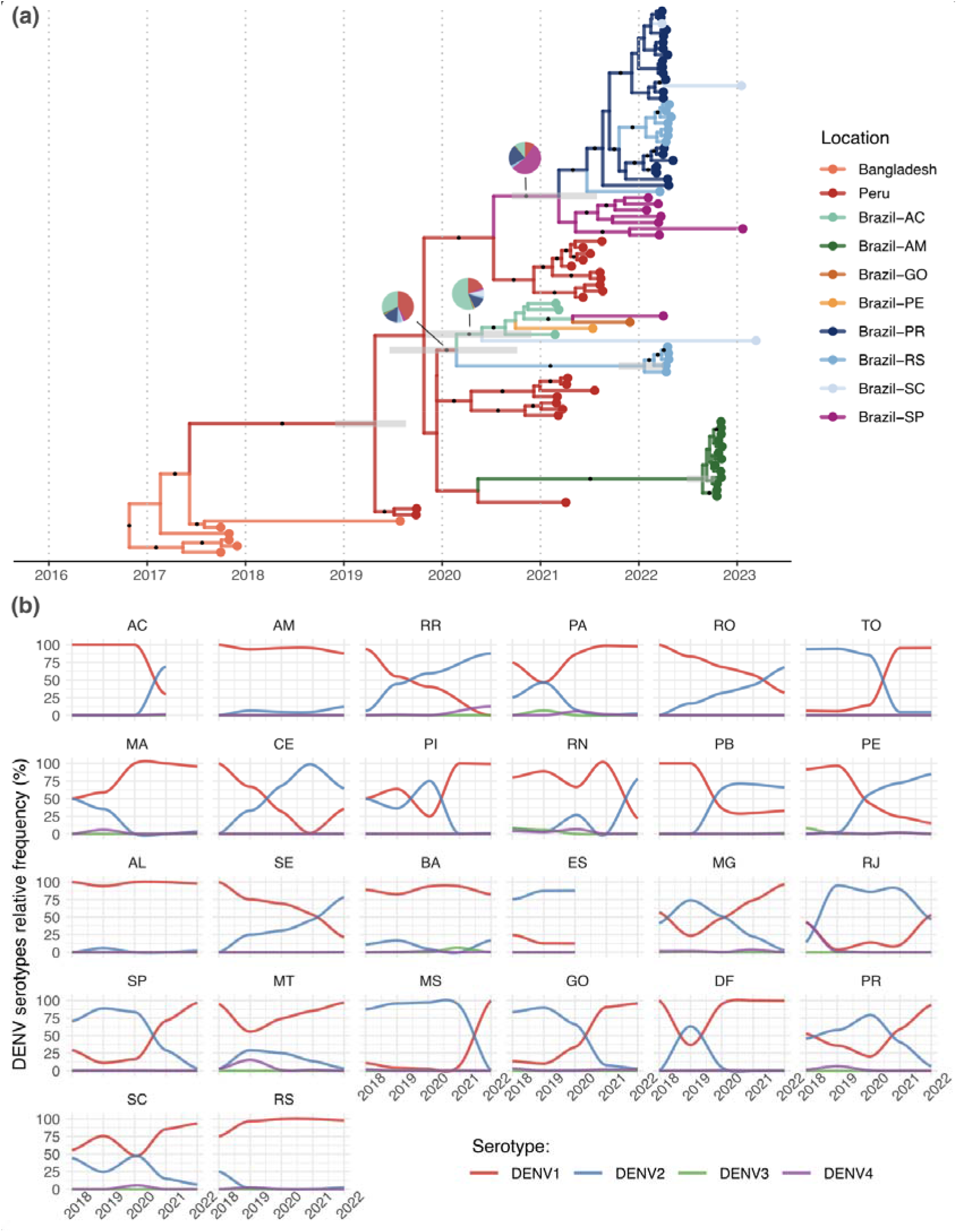
Spatio-temporal evolution of DENV-2 genotype II (Cosmopolitan) in South America and circulation dynamics of DENV serotypes in Brazil in recent years. (a) Time-scaled phylogenetic tree with ancestral locations inferred by the phylogeographic model. Branches are colored according to the most likely ancestral location, and pie-charts show the posterior probability of each location for key nodes with high uncertainty. Error bars show the 95% high posterior density of node heights related to introductions to Brazil. Black dots on branches indicate a posterior probability higher than 0.9. (b) Relative frequency of each DENV serotype in all Brazilian states between 2018 and 2022. States acronyms are according to the ISO 3166-2:BR.

To assess the potential of DENV-2-GII to spread and trigger relevant outbreaks in Brazil, we analyzed the dynamics of DENV serotypes’ circulation in the country as identified through PCR tests by the laboratory surveillance and informed in the national system of diseases of compulsory notification (SINAN). Between 2018 and 2022, the Brazilian dengue epidemic was dominated by DENV-1 and DENV-2, with significant regional differences (**Figure 1B**). A predominance of DENV-2 after 2020, which could indicate a genotype II outbreak, was observed in the Northern states of AC (where genomic surveillance detected DENV-2-GII circulation), RO and RR, in the Southeastern states of RJ and Espirito Santo (ES), and in the Northeastern states of Ceara (CE), Paraíba (PB), Rio Grande do Norte (RN), Sergipe (SE) and PE. In the latter, only one sequence of DENV-2-GII was found compared to 80 sequences of DENV-2-GIII in 2021, indicating that the dominance of DENV-2 in PE at that time was not caused by genotype II, which could also be the case for the neighboring Northeastern states. A high prevalence of DENV-2 was observed in several states between 2018 and 2020, but there was no evidence of circulation of DENV-2-GII in Brazil before 2021 (**Supplementary Figure 2**). Finally, in other states including AM and RS, where genomic surveillance detected recent introductions of DENV-2-GII in 2022, DENV-1 was responsible for most dengue cases in the whole period supporting that DENV-2-GII only caused localized outbreaks in those states. In fact, in 2022, DENV-1 was responsible for more than 80% of the dengue cases in 17 out of 25 Brazilian states with data available.

In summary, this study confirms that DENV-2-GII is circulating nationally in Brazil, expanding the findings of previous localized studies^6,7^. Our phylogenetic analysis unveiled multiple introductions of this genotype into Brazil from Peru, generating multiple clades that spread in the country with different success. Despite the wide geographic dispersion of DENV-2-GII, dengue epidemics in 2021-2022 in most Brazilian states were dominated by DENV-1, supporting that circulation of DENV-2-GIII in previous years probably created a high population immunity barrier against transmission of DENV-2-GII in Brazil. However, the rapid spread of DENV-2-GII across the country’s territory and the heterogeneous population immune landscape across regions, could create an epidemic scenario for sustained cryptic circulation and localized outbreaks. Indeed, DENV-2-GII was probably responsible for most DENV-2 infections in Brazil in 2022 and this warrants close monitoring to better understand the potential impact of this genotype in the coming years.

## Supporting information

Supplementary Consortia List

## Data Availability

All data produced in the present study are available upon reasonable request to the authors

