## Supplementary text for "Multiple introductions and country-wide spread of DENV-2 genotype II (Cosmopolitan) in Brazil"

### Materials and Methods

#### Ethics statement and samples

This study was approved by the Ethics Committee of Instituto Oswaldo Cruz, which waived signed informed consent (CAAE: 90249218.6.1001.5248). A collaborative network was established with the Central Laboratories from Brazilian states of Amazonas (LACEN-AM), Pernambuco (LACEN-PE), São Paulo (LACEN-SP/Instituto Adolfo Lutz [IAL]), Paraná (LACEN-PR), Santa Catarina (LACEN-SC) and Rio grande do Sul (LACEN-RS).

LACENs tested arboviral suspected cases using a reverse transcriptase real-time PCR kit that detects Zika, dengue, and chikungunya, and identifies the four dengue serotypes (IBMP - Kit Biomol ZDC - https://www.ibmp.org.br). A selection of confirmed cases of dengue serotype 2 (DENV-2) infection, sampled between March 2021 and March 2023, were then submitted to viral whole-genome sequencing.

#### Whole-genome sequencing and genome assembling

Sequencing was performed in Oswaldo Cruz Foundation (Fiocruz) laboratories participating in the Fiocruz Genomics network (https://www.genomahcov.fiocruz.br/en/), in IAL and LACEN-RS. Selected samples were submitted to viral total nucleic acid extraction and used in sequencing library preparation with Illumina's Viral Surveillance Panel (VSP w ILMN RNA Prep w Enrich) or Illumina's COVIDseq assay adapted to DENV sequencing. Briefly, DENV-2 specific primers were designed to amplify the whole viral genome using a tilling amplicon strategy[1] . Primers were designed to maximize positions of conserved regions based on the available Brazilian genomic sequences downloaded from the ViPR database (https://www.bv-brc.org/) on April 2018. The final set of primers including degenerated bases generate 425 bp amplicons with 50 bp of overlap and were designed to amplify the Asian-II, Asian-I, Asian-American and Cosmopolitan DENV-2 genotypes. Primers' sequences can be found in **Supplementary Table 2**. DENV-2 primers were then used in substitution to SARS-CoV-2 primers and CONVIDseq library preparation was performed according to manufacturer's instructions.

Libraries were then submitted to nucleotide sequencing on a MiSeq instrument (Illumina). The FastQ files generated at Illumina's cloud (https://basespace.illumina.com) were downloaded and quality control, trimming and mapping against DENV-2 reference was performed in ViralFlow dev_1.0 version [2] . Genomes generated with Illumina's VSP were assembled with a customized workflow using Geneious Prime 2022.

#### DENV-2 genotype identification and global sequence dataset assembling

The DENV-2 consensus genomes obtained in this study were initially submitted to genotype identification using the Flavivirus Genotyping Tool Version 0.1 (https://www.rivm.nl/mpf/typingtool/flavivirus/) or Dengue Virus Typing Tool at Genome Detective (https://www.genomedetective.com). The DENV-2 genotype II genomes identified here were then aligned to a global dataset composed by all DENV-2 sequences with more than 50% of coverage breadth and with complete data for location and date of sampling, as available in NCBI Virus (https://www.ncbi.nlm.nih.gov/labs/virus/vssi/#/) on 10th May, 2023. Sequences were aligned with nextalign [3] and to identify the global genotype II clade, a maximum likelihood (ML) phylogenetic tree was inferred with IQ-TREE [4] using the fast mode and GTR+G4 substitution model. Sequences belonging to genotype II were then submitted to a second IQ-TREE run using the standard strategy of tree search, GTR+G4 substitution model and branch support was inferred with SH-aLRT on 1000 replicates.

#### Spatiotemporal evolutionary history of DENV-2 genotype II in Brazil

To reconstruct the spatiotemporal history of DENV-2 genotype II in Brazil, the whole American clade, including basal sequences from Bangladesh, as identified in the ML tree, were submitted to a Bayesian phylogeographic inference approach using BEAST v1.10.4[5] . Using TempEst v.1.5.3[6] , outlier sequences that deviated more than 3.0 interquartile ranges in the regression analysis of the root-to-tip divergence against the tip sampling time in the ML phylogenetic tree of DENV-2 genotype 2 were removed. Time-scaled trees were inferred with a relaxed molecular clock model, which outperformed the strict clock model in marginal likelihood estimation, and a Bayesian Skyline coalescent model. The ancestral node states were reconstructed with a continuous-time Markov chain (CTMC) prior and discrete spatial diffusion with an asymmetric substitution model to infer the migration events. Bayesian stochastic search variable selection (BSSVS) was performed to identify the significant migration routes [7]. Markov Chain Monte Carlo (MCMC) was run for 100 million generations and convergence was assessed by calculating the Effective Sample Size (ESS) for all parameters using Tracer v1.7.1 [8]. Maximum clade credibility (MCC) tree was summarized with the TreeAnnotator tool (part of the BEAST package), imported into RStudio with “treeio” [9] and plotted with “ggtree” [10] package.

#### Epidemiological data collection and visualization

Epidemiological data on dengue fever cases in Brazil were downloaded from the DATASUS database using the R package microdatasus [11] . Specifically, data on the number of confirmed DENV cases and their corresponding serotypes were obtained from 2018 to 2022. The data were extracted at the national level and stratified by year and state. Plots were produced using the ggplot2 package on R software version 4.1.2.

#### Data availability

All DENV-2 genotype II sequences generated in this study were made available in GenBank and EpiArbo at GISAID (**Supplementary Table 1**) .


#### Acknowledgments

We acknowledge all data contributors, i.e., authors and their originating laboratories responsible for obtaining and timely submitting dengue virus genomes, including metadata, via GenBank or other public databases. We appreciate the support of FIOCRUZ Genomics Surveillance Network members. Funding support: FAPEAM (Universal/AM call 2019; Rede Genômica de Vigilância em Saúde - REGESAM); Inova Fiocruz/Fundação Oswaldo Cruz (Inova Amazônia); Departamento de Ciência e Tecnologia (DECIT) of the Brazilian MoH; G.B. is supported by CNPq through a productivity research fellowships (304883/2020-4) and FAPERJ (Grant number E-26/202.896/2018).
