## Supplementary Figure 1 for "Multiple introductions and country-wide spread of DENV-2 genotype II (Cosmopolitan) in Brazil"

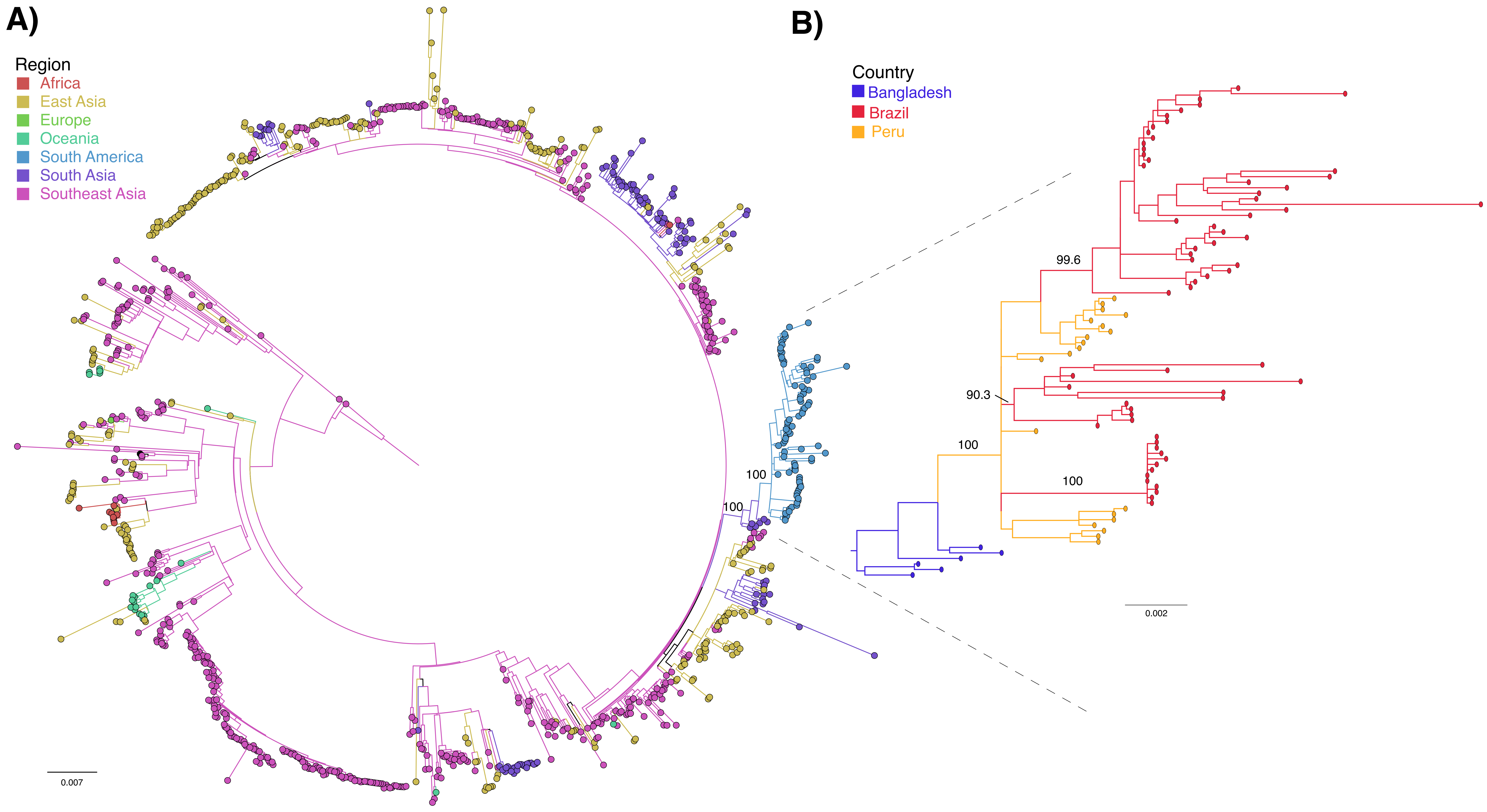


**Supplementary Figure 1. Maximum phylogenetic (ML) tree of the global diversity of DENV-2, genotype II and identified South American clade.** (a) Circular ML tree representing DENV-2, genotype II, diversity found in the world and colored according to relevant regions. (b) DENV-2, genotype II, South American clade colored according to the country of sampling and basal sequences collected in Bangladesh. Branch support for important clades are shown.
