## Supplementary Figure 2 for "Multiple introductions and country-wide spread of DENV-2 genotype II (Cosmopolitan) in Brazil"

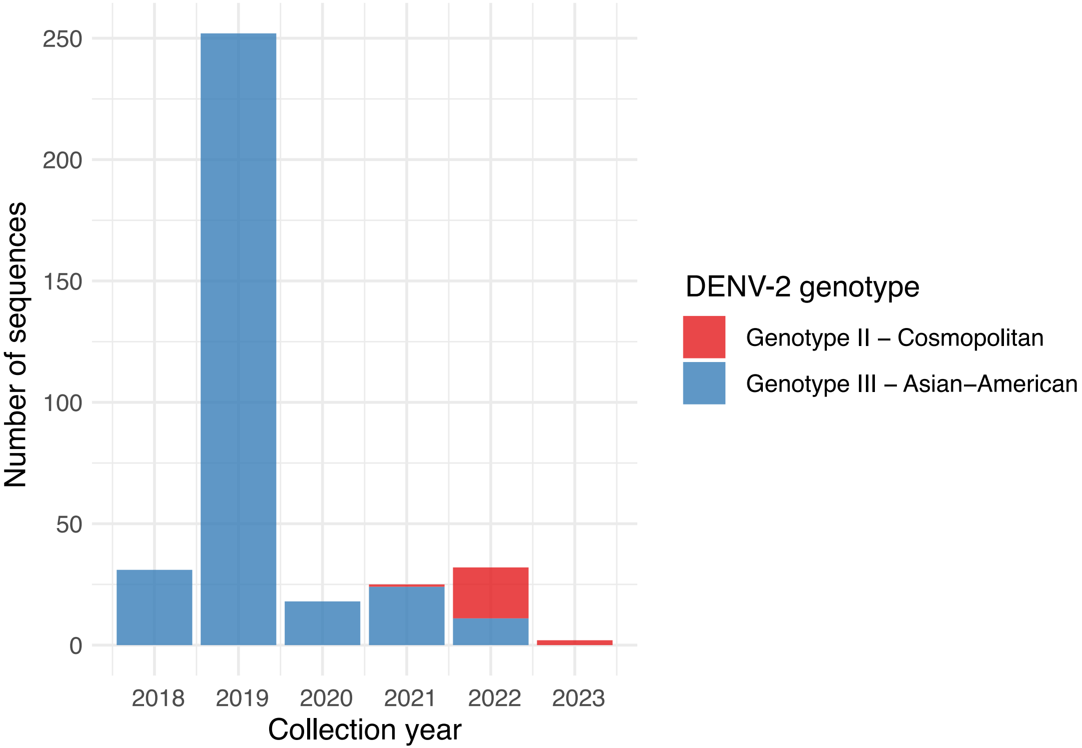


**Supplementary Figure 2. DENV-2 genotype diversity circulating in Brazil between 2018 and 2023.** All DENV-2 sequences from Brazil, as available at NCBI virus on 10th May, 2023, were classified into genotypes with online tools and collection date was aggregated in years. DENV-2 genotype is colored according to the legend.
